# Alpha band oscillations in the limbic pallidum track future alcohol use

**DOI:** 10.1101/2025.07.14.25331334

**Authors:** Louis Sauter, Kelly Kadlec, Collin Lehmann, Sharvari Shivanekar, Payel Roy, Donald Crammond, Brian Coffman, Jorge González Martínez, Khaled Moussawi

## Abstract

Alcohol use disorder (AUD) is difficult to treat, with high relapse rates even after prolonged abstinence. The limbic pallidum (LP) is a key structure involved in reward and motivated behavior and has been proposed as a potential target for neuromodulatory interventions for AUD like deep brain stimulation (DBS). In this study, a sensing-enabled DBS system was implanted in the LP of a patient with severe, treatment-resistant AUD. Local field potentials (LFPs) were recorded during regular clinic visits and cue reactivity sessions to examine whether neural activity in the LP is related to alcohol use or alcohol craving. We identified prominent neural oscillations in the canonical alpha to low beta frequency range (7-15 Hz) that peaked in the alpha band (11-12Hz) and was localized to the right ventral LP. Relative power across this frequency range was strongly correlated with the number of drinks consumed in the days following the recording session, with the strongest correlation observed with drinking behavior in the first three days after the recording session. Furthermore, the heavy vs. light or no drinking behavior could be distinguished based on the power of the LP alpha oscillation using unbiased clustering. The correlation was specific to the right ventral LP and was only significant for drinking behavior following, rather than preceding, recording sessions. These results suggest that LP oscillations in the alpha band may serve as a biomarker of future heavy alcohol use and highlight the possibility of identifying a neural biomarker of drinking behavior that could be used to guide AUD treatment.

## 1. Introduction

The currently available therapies for alcohol use disorder (AUD) have limited efficacy, with an estimated 40-60% of treated patients relapsing after abstinence within 6 months (Anton, O’Malley et al. 2006). This, paired with the devastating societal and economic cost of AUD, has motivated extensive research into the neural circuit dysfunctions of AUD, highlighting several brain networks of importance (Koob & Volkow, 2016). The limbic pallidum (LP) is a critical node at the convergence of the different circuits implicated in AUD pathophysiology and is thought to be a promising therapeutic neuromodulation target for AUD (Prasad and McNally 2020).

Studies in monkeys and humans are consistent with rodent studies regarding the LP’s role in reward and aversion processing (Bhatia and Marsden 1994, Pessiglione, Schmidt et al. 2007, Childress, Ehrman et al. 2008, Cilia, Siri et al. 2008, Napier and Mickiewicz 2010, Tachibana and Hikosaka 2012, Howell, Prescott et al. 2016, Saga, Richard et al. 2016, Saga, Ruff et al. 2019). In humans, the LP includes the ventral pallidum (ventral to the anterior commissure) and anteromedial globus pallidus (internal and external segments) (Haber and Knutson 2010). The LP receives dense input from the ventral striatum, amygdala, and hippocampus, and projects to the thalamus from which thalamocortical efferents project widely to many cortical areas governing affect and reinforcement, positioning it as a major integrator of reward-related information to regulate motivated behavior (Prasad and McNally 2020). The preclinical literature shows that GABAergic neurons in the LP encode rewards and drive approach behaviors, and that the LP is critical for drug seeking and relapse in various substance use disorders – including alcohol, stimulants, and opioids – and is essential for resumption of drug seeking after abstinence (Farrell, Ruiz et al. 2019, Prasad and McNally 2020). On the other hand, the LP glutamatergic neurons encode aversive outcomes and drive avoidance behavioral responses (Prasad and McNally 2020).

The Percept DBS neurostimulator from Medtronic allows for recording of local field potentials (LFPs) from implanted structures and is increasingly used to study the neural basis of pathological behaviors in various neuropsychiatric disorders, including Parkinson’s disease, obsessive compulsive disorder, depression, and addiction (Groiss, Wojtecki et al. 2009, Lavano, Guzzi et al. 2018, Alagapan, Choi et al. 2023, Vissani, Nanda et al. 2023). Furthermore, symptom-based titrations and drug cue-reactive electrophysiological biomarkers have been used to personalize and target opioid use disorder and AUD (Kuhn, Moller et al. 2014, Mahoney, Haut et al. 2021, Davidson, Giacobbe et al. 2022, Bach, Luderer et al. 2023, Qiu, Nho et al.2025). We have implanted the Percept DBS system and Sensight electrodes (Medtronic Inc.) into the LP in a patient with severe AUD and alcohol associated liver disease. We recorded LFPs from the LP during clinic visits and measured changes in LFP oscillatory activity within specific frequency bands using aperiodic fitting tools. Given the role of the LP in reward and motivated behavior and in relapse to alcohol seeking, we hypothesized that neural activity in the LP, recorded as LFPs, could be associated with alcohol-related behaviors (e.g., consumption, craving).

## 2. Methodology

### 2.1 Subject, Surgery, and Device

A male in his 50s with severe and refractory AUD and advanced compensated liver disease (fibrosis stage F3/F4) was implanted with Sensight DBS electrodes (Medtronic Inc.) into the limbic pallidum (anterior GPi – Globus Pallidus internal segment). A Percept neurostimulator (Medtronic Inc.) was implanted in the right anterior chest wall. The study (NCT05522751) complies and was approved by the University of Pittsburgh’s Institutional Review Board. This study was designed as an open-label study to evaluate the safety and feasibility of limbic pallidum DBS for the treatment of severe and refractory alcohol use disorder (AUD) with concomitant advanced, asymptomatic alcoholic liver disease (ALD). AUD severity was determined by meeting >6 DSM-5 criteria (American Psychiatric 2013).

Per the study protocol, after the informed consent process, the subject underwent baseline evaluations that involved assessment of alcohol use and craving. The DBS system was implanted within a month from the baseline assessments and turned on and optimized 4 weeks after the surgery (stimulation frequency ∼125 Hz; pulse width 90 μs pulse; current intensity of 3-5 mA).The DBS electrodes were localized using the LeadDBS software (Horn, Li et al. 2019). The subject was followed with weekly phone visits to ensure safety and accurate alcohol use reporting with Timeline Follow Back (TLFB), and monthly in-person visits during which LFPs were recorded in indefinite streaming mode for 10-20 min. During each visit, the participant underwent clinical assessments including Alcohol Craving Questionnaire-short (ACQ) (Browne, Wray et al. 2016). The clinical evaluations and outcomes of DBS treatment will be reported in a separate manuscript. There were 16 recording sessions, 8 CR sessions, and 12 ACQ surveys administered. Some recording sessions did not have corresponding CR and ACQ scores and vice versa. A breathalyzer test was administered at the beginning of each session to assess for acute intoxication.

### 2.2 Cue Reactivity

During eight visits, the participant underwent a cue reactivity task. Each task session lasted approximately 12 minutes, comprising eight blocks of personalized alcohol-related (Drink) or neutral pictures. Each block consisted of 5 pictures presented in succession, each for 5 s (25 s total). The pictures were then followed by a prompt to rate craving, positive and negative affect, calm, and excitement using a visual analog scale (VAS, 0-100) presented on a monitor with a mouse slider. The blocks were presented in a fixed order between sessions: Neutral, Drink, Neutral, Neutral, Drink, Drink, Neutral, Drink. Baseline DBS-LFPs were acquired 2-5 minutes before each cue reactivity task. A timer synchronized with the start of LFP recording was used to mark the onset and offset of each cue reactivity block, including transitions between Drink and Neutral conditions, picture presentations, and VAS question prompts.

### 2.3 Data Processing and Analysis

LFPs were recorded in bipolar configuration between three unique pairs of contacts in each lead (0-3, 1-3, and 0-2 in the left hemisphere, and 8-11, 9-11, and 8-10 in the right hemisphere) in the Percept’s indefinite streaming mode (stimulation off). Electric potentials between adjacent contacts (0-1, 1-2, and 2-3 on the left, and 8-9, 9-10, and 10-11 on the right) were computed from the raw LFP recordings. LFPs were divided into 3-second epochs, and each epoch was windowed and passed to MATLAB’s periodogram function. The periodogram computes the power spectral density by taking the squared magnitude of the Fast Fourier Transform (FFT) of the windowed signal and normalizing by the length of the epoch. The calculated power spectral densities (PSDs) were then averaged for each recording session. Periodic spectral power was separated from the 1/f aperiodic component of the PSDs with a fitting of one over frequency (FOOOF) algorithm (Donoghue, Haller et al. 2020, Bush, Zou et al. 2023). Fitting was done with a 50% threshold for including datapoints from 5 to 50 Hz in the preliminary aperiodic fit, 5 max periodic peaks to detect, and peak detection thresholds of either 0.38 standard deviations or 0.11 decibels. The algorithm attempted to include a knee parameter for the aperiodic fit. Post-fitting corrections were made if the minimum power difference between the first 20 datapoints of the fit and the spectra was above the power threshold (0.11 decibels) for more accurate comparisons of spectral peak heights. To compare frequency band power, relative periodic power (RPP) was calculated by integrating the periodic PSD over specific frequency ranges (theta: 5-8 Hz; alpha: 8-12 Hz; low beta: 13-20 Hz; high beta: 21-30 Hz; low gamma: 31-50 Hz; high gamma: 51-125 Hz). The observed prominent oscillations occurred within the 7-15 Hz range, with a peak in the alpha band (11-12 Hz). Given this observed oscillation range that extends beyond the canonical alpha band, which is historically defined based on cortical oscillations, and given the evidence that cortical and subcortical oscillations do not necessarily align perfectly (Brown and Williams 2005, Lalo, Thobois et al. 2008, Frauscher, von Ellenrieder et al. 2018), we refer to the observed LP alpha band activity as the frequency band of interest (FBOI). The computed PSDs were also compared to the FFT magnitude derived from the Percept in survey mode to confirm consistent trends in the data. Bivariate relationships between RPP and alcohol craving, alcohol use, and other behaviors were assessed using Spearman correlation. RPP in different frequency bands from heavy drinking and no/light drinking – as defined by the National Institute on Alcohol Abuse and Alcoholism (heavy drinking in men: >5 drinks/day or >15 drinks/week) – were compared using 2-way Repeated Measures ANOVA. The aperiodic fit parameters were also split into the same groups and compared using the Mann-Whitney U test. The Dunn-Šidák test was used to control for multiple comparisons where appropriate.

## 3. Results

### 3.1 Alpha to low beta power is strongly correlated with future alcohol use

PSDs were generated from 16 recording sessions, separated into periodic and aperiodic components, and averaged across all sessions. The frequency domain data revealed prominent oscillatory activity in the 7-15 Hz range that peaks in the alpha band (11-12 Hz), which was recorded from the most ventral contacts of the LP DBS electrodes in the right hemisphere (8-9) (Fig 1a, b). We refer to the 7-15 Hz frequency range as the frequency band of interest (henceforth FBOI), given that it extends beyond the canonical alpha band (8-12Hz), as described in the methods section. A distinct peak in the alpha/FBOI band was not observed in recordings from other more dorsal contacts on the right, or any contacts on the left, but was observed between contact pairs 8-10 and 8-11, suggesting its specificity to contact 8 (Fig S1a). A second prominent peak in the high beta range (21-30 Hz) was observed between right-sided contacts, most pronounced between contacts 9 and 10 sequentially (Fig 1b), and contacts 8 and 11 overall (Fig S1a).

**Figure 1.**
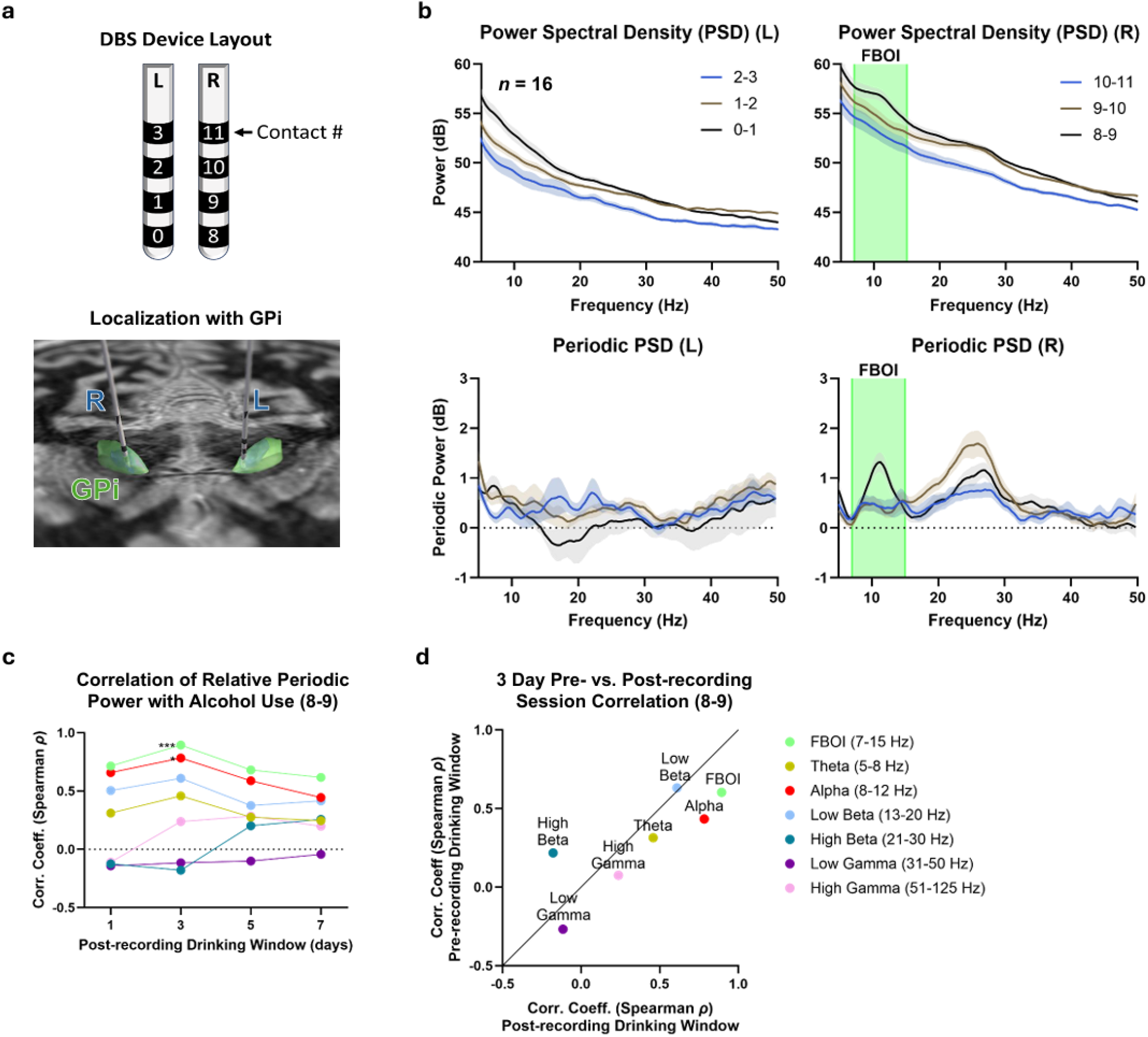
Alpha to low beta power is strongly correlated with future alcohol use. **(a)** Layout and localization of the DBS electrodes, with ‘L’ and ‘R’ denoting the left and right hemispheres of the brain, respectively, and the 4 contacts on each numbered ventral to dorsal. The electrodes are localized to the anterior limbic pallidum. The Globus Pallidus internal segment (GPi) is shown in green. Electrode placement is asymmetric, with the right electrode more ventral. **(b)** Top: mean power spectral densities (PSDs) between each sequential contact pair from all recordings (*n* = 16) with standard error of the mean (translucent shading). Green shading highlights the frequency band of interest (FBOI, 7-15 Hz). Bottom: isolated periodic component resulting from fitting of one over frequency (FOOOF) for each PSD. **(c)** Spearman’s rank correlation coefficients for relative periodic powers (RPPs) from contact pair 8-9 correlated to total number of drinks post-recording sessions. The drinking window refers to the number of days from the day of the recording that were used to determine the total number of drinks within that window (Šidák’s multiple comparison test; \*\*\**p* = 0.0002, \**p* = 0.0194). **(d)** Spearman’s rank correlation coefficient over a 3-day drinking window for pre- and post-recording for all investigated frequency bands.

We examined the relationship between craving scores and spectral features of the recorded LFPs. We found no significant correlation between craving and relative periodic power (RPP) of canonical frequency bands (Fig S2b). Differences in Alcohol Craving Questionnaire (ACQ), alcohol craving during the CR task, and differential craving scores (alcohol vs neutral blocks in the CR task) did not fluctuate much from zero and therefore the correlations were not meaningful (Fig S2d). Breathalyzer tests for acute intoxication returned negative on 77% (10/13) of sessions.

The neural oscillatory activity in the alpha to low beta band (FBOI) was significantly correlated with drinking behavior. To explore the relationship between the recorded LFPs and drinking behavior, we sought to examine the correlation between drinking behavior, measured as the number of drinks, before, after, and around each recording session (Fig S3a), and the spectral power in various frequency bands. The RPP was integrated for each of the canonical frequency bands plus the FBOI. The Spearman correlation coefficient between the resulting band powers and number of drinks consumed in the 1, 3, 5, and 7 days prior, post, and around each recording session were computed (Fig S3b). The greatest correlation was observed between the FBOI RPP and the number of drinks consumed in the 3-days following the recording session (Fig 1c, Spearman’s *ρ* = 0.8935, Šidák’s multiple comparison test; *p* = 0.0002). This correlation was stronger for future drinking compared to past drinking (Fig 1d), and was limited to the FBOI and its subcomponent, canonical alpha band. Excluding sessions with positive breathalyzer test did not alter the strength of the correlation (Spearman’s *ρ* = 0.8838; *p* = 0.0001).

Other time windows analyzed along with aperiodic fit parameters did not exhibit strong correlations (Fig S3, S4). Similar analyses for other contacts did not reveal meaningful correlations (Fig S4). Of note, the FBOI RPP did not show significant correlation with depression (MADRS) and anxiety (HAM-A) scores (Fig S2c).

### 3.2 Periodic Activity in Heavy vs. Light Drinking

Automatic clustering with the K-means method (*k* = 2) of the FBOI periodic power and number of drinks (3 days post recoding) resulted in two clusters that aligned with heavy (*n* = 7 datapoints) and no/light drinking (*n* = 9) as defined by the National Institute on Alcohol Abuse and Alcoholism (heavy drinking in men: >5 drinks/day or >15 drinks/week) (Fig 2a). Therefore, we separated the recording sessions based on no/light and heavy drinking (Fig 2b). The periodic FBOI power observed between contacts 8 and 9 was significantly greater in heavy compared to no/light drinking (Fig 2c, d *p* < 0.001). Alpha band periodic power was also significantly different between contacts 8 and 9 (Fig 2d, *p* < 0.05). No other significant differences were observed in either RPP in heavy vs. no/light drinking between other sequential contact pair (Fig S5). Further, no differences were observed between the fitting parameters for the aperiodic component of the power spectra (Fig 2e, S5).

**Figure 2.**
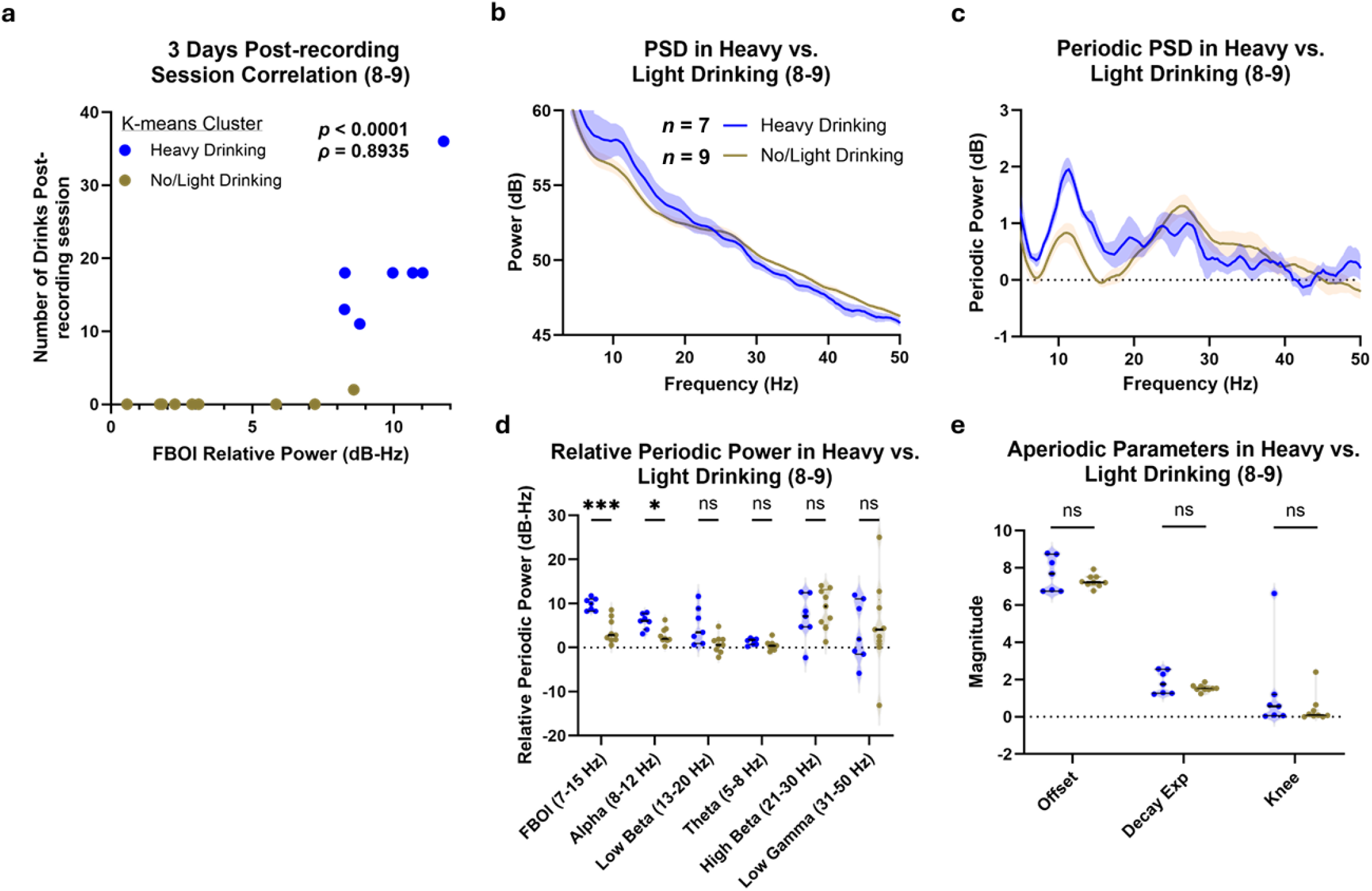
PSD in the FBOI tracks heavy vs. light future drinking behavior. **(a)** Scatterplot showing the strongest correlation, relative periodic power (RPP) from contact pair 8-9 across FBOI and number of drinks 3 days following the recording session (Spearman’s *ρ* = 0.8935, *p* < 0.0001). Points colored by result of k-means clustering (*k* = 2, using Euclidian distance) into heavy drinking (blue) and no/light drinking (brown) recording sessions. **(b)** Mean power spectral densities (PSDs) with standard error of the mean (translucent shading) grouped by heavy alcohol consumption following recording (blue, *n* = 7) or no/light alcohol consumption following recording (brown, *n* = 9). **(c)** Periodic component of PSDs resulting from fitting of one over frequency (FOOOF) for each recording. **(d)** RPP in heavy drinking (blue) vs. no/light drinking (brown), ordered left to right from largest to smallest effect (2-way repeated measures ANOVA; Heavy vs. No/Light Drinking effect *F*(1, 14) = 4.629, *p* = 0.0494, Šidák’s multiple comparison test; \*\*\**p* = 0.0005, \**p* = 0.0214). **(e)** Aperiodic fit parameters compared in heavy drinking (blue) vs. no/light drinking (brown) (Mann-Whitney U test).

### 3.3 Effect of Cue Presentation on Periodic Activity

LFPs were recorded before and throughout the cue reactivity task during select visits (*n* = 8). Power spectra were computed for pre-task baseline and cue-presentation periods and decomposed into periodic and aperiodic components by FOOOF. Periodic components were averaged across sessions for baseline and cue periods. Across all cue reactivity sessions (neutral and drink cues), the average PSD power in the FBOI band was not statistically different during the presentation of alcohol images, neutral images, and baseline (Fig 3a, b). We did not observe differences in the PSD power during alcohol vs. neutral cue presentations for any spectral band. However, we observed a significant difference between drink cue presentation vs. baseline in the low beta band (Fig 3b, *p* = 0.0321). When sessions were separated into heavy and light drinking as previously defined (Fig 2a), the FBOI periodic power increased with cue presentation compared to baseline in the light drinking group but not the heavy drinking group (Fig 3c).However, the number of sessions was low for a meaningful statistical comparison.

**Figure 3.**
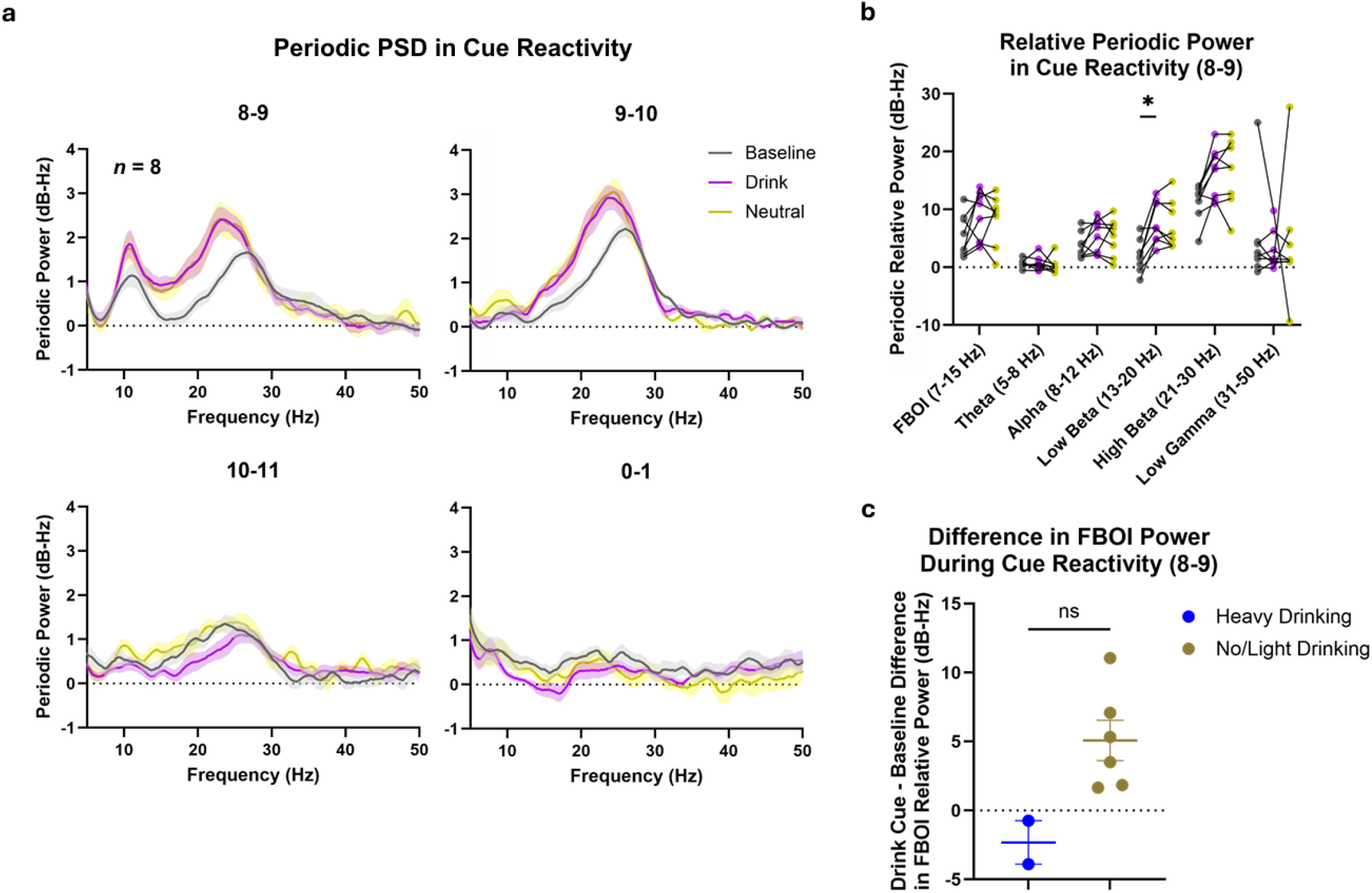
Spectral power response to cue presentation differs based on drinking behavior. **(a)** Isolated periodic component resulting from fitting of one over frequency (FOOOF) from recorded LFPs during baseline (black) and cue presentations (drink in purple and neutral in yellow) (*n* = 8), from various electrode contacts.**(b)** Integrated relative periodic power (RPP) during alcohol and neutral cue presentations relative to baseline (2-way repeated measures ANOVA, Šidák’s multiple comparison test; \**p* = 0.0321). **(c)** FBOI power in heavy drinking (blue) vs. no/light drinking (brown) during cue reactivity with standard error of the mean shown (Mann-Whitney U test).

## 4. Discussion

In this exploratory, hypothesis-generating study, we identify a potential neural biomarker of future alcohol use. In a patient with severe alcohol use disorder (AUD), increased oscillatory activity in the alpha to low beta range in the right limbic pallidum (LP) was strongly correlated with alcohol consumption over the following days, which was strongest at 3 days post-recording session.

The first key aspect of our finding is the anatomical specificity of the signal. The increased FBOI periodic activity was most strongly observed between contact pair 8-9, which corresponds to the right ventral pallidum, ventral to the GPi. The increased alpha band activity was also recorded between contact pairs 8-10 and 8-11, but not contact pairs 9-10 and 10-11 on the right, or any contact pairs in the left hemisphere. This localizes the activity to contact 8, which was in the ventral pallidum, ventral to the anterior GPi. Recently, a similar contact-level localization was reported in opioid use disorder, where cue reactivity oscillations within a single Nucleus accumbens contact pair guided DBS targeting (Qiu, Nho et al. 2025). Together, these findings indicate that discrete subregions of the basal ganglia harbor electrophysiologic signatures predictive of substance use, strengthening the rationale for circuit-specific biomarkers in closed-loop neuromodulation. However, our results do not suggest that this signal is unilateral. This is because the electrodes were not symmetrically implanted between the two hemispheres, as the most ventral contact of the left hemisphere was within the ventral GPi and is not equivalent to contact 8 on the right.

Alcohol use is sometimes co-morbid with essential tremor or alcohol withdrawal tremor, both characterized by muscle activity in the theta and alpha range (Schroeder and Nasrallah 1982, Clark and Louis 2018). Essential tremor has been shown to be associated with neural oscillatory activity in the 4-6 Hz range in the motor basal ganglia (Hutchison, Lozano et al. 1997). Thus, it is theoretically possible that the observed LP alpha oscillation could be partially confounded by tremor activity. However, the study patient’s neurological examination during each visit did not reveal any signs of essential tremor or visible alcohol tremor. Further the electrode contact 8 was located far anterior, medial, and ventral, away from the motor GPi where alpha oscillations related to tremor are usually recorded. Finally, alcohol withdrawal tremor is usually bilateral, so we would expect bilateral alpha oscillations in the GPi, but this was not observed here.

Therefore, it is unlikely that the unilateral right alpha range oscillations we observed from the ventral pallidum were encoded tremor activity of the left hand. In future studies, the possibility of a tremor confound could be ruled out with simultaneous limb electromyography (EMG) recordings.

The electrophysiological recordings were mainly obtained with documented negative breathalyzer testing, and the alpha band/FBOI RPP was more correlated with future drinking than past drinking – most strongly correlated with 1-3 days future drinking. These results suggest that the observed increase in the alpha and low beta power is not simply due to acute effects of alcohol on physiological activity, or acute neurophysiological adaptations in response to alcohol exposure. In such cases, we would expect the power to be more correlated with drinking prior to the recording session. Past research demonstrates how medial prefrontal cortex activity in response to persuasive health messages can predict behavior changes in the following week without self-reported attitude or intention changes (Falk, Berkman et al. 2010). Our results express similar findings, where self-reported craving scores are unrelated to drinking behavior and, instead, specific neural activity can better predict drinking behavior changes.

Interestingly, we found that the alpha band/FBOI periodic activity associated with cue reactivity did not always increase during drug cue presentation. Instead, this RPP only increased during cue-reactivity sessions followed by no or light drinking, which could suggest a peak effect of the LP FBOI/alpha PSD power (power increases in response to craving only when it is low to start with but cannot increase when it is already high). Notably, baseline drug craving has been reported to have no significant impact on the effectiveness of cue reactivity to cause increases in drug use and craving (Shiffman, Shadel et al. 2003, Janes, Krantz et al. 2020).

We did not observe a correlation of the FBOI RPP with craving scores. While spontaneous craving or craving in response to cue presentation (as in the cue reactivity task) may generally correlate with and predict alcohol use (Witteman, Post et al. 2015, McHugh, Fitzmaurice et al. 2016), the craving reports from our study subject did not vary much and did not correlate or predict his alcohol use. This highlights the limitation of using the craving construct as it relies on subjective report.

We also found that cue exposure – computer screen presentation of pictures containing either alcohol or neutral cues – increased low beta band periodic activity for each recording, with no correlation to alcohol consumption. Previous work has shown that higher levels of visual attention are associated with increased beta power and decreased alpha (Hanslmayr, Aslan et al. 2007), which could explain the increases in beta power and variability of alpha power during cue presentation. Differences in drink vs. neutral cue presentation were insignificant, most likely due to limitations in synchronizing the LFP timing with the different blocks in the CR task, which could have confounded our results.

Other potential influences on the periodic alpha wave activity in the LP are negative mood states (Neumann, Huebl et al. 2014). Neurons in the LP are responsible for wakefulness and various emotions such as depression and anxiety (Luo, Ge et al. 2023), and major-depressive disorder, anxiety disorders, and stress-related disorders are often associated with AUD (Prasad and McNally 2020). However, we did not observe a correlation between the alpha band or FBOI RPP and depression and anxiety scores.

The primary limitation of this study is the *N* of 1. Replication in other patients is necessary to confirm the reliability and generalizability of the findings. Additionally, the results reported here are mainly correlational. Suppressing alpha band RPP in the LP with DBS is one potential way to assess the causal relationship of this RPP and drinking.

In summary, our findings suggest an increase in periodic activity across the canonical alpha band which extends into the low beta range in the LP as a marker of future heavy drinking. Since craving is inherently subjective, there is a need to identify related electrophysiological biomarkers for AUD. Future experiments involving larger and more diverse subject populations are warranted to investigate periodic activity across the alpha and other bands as a biomarker for craving.

## Data Availability

All data produced in the present study are available upon reasonable request to the authors

