## Supplemental Figures for "Alpha band oscillations in the limbic pallidum track future alcohol use"

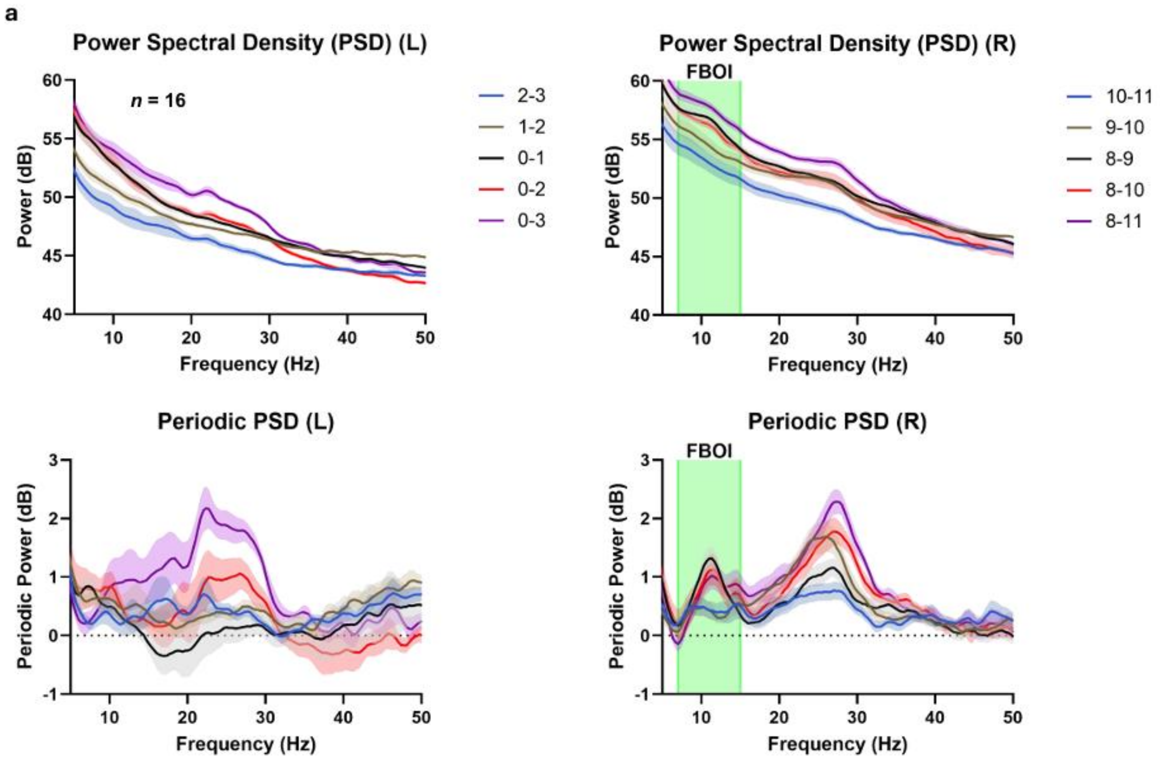


**Supplemental Figure 1. Non-sequential contact power spectral densities emphasize specificity of the FBOI to contact 8. (a)** Top: mean power spectral densities (PSDs) between each sequential contact pair and two non-sequential contact pairs from all recordings (*n* = 16) with standard error of the mean (translucent shading). Green shading highlights the frequency band of interest (FBOI, 7-15 Hz). Bottom: isolated periodic component resulting from fitting of one over frequency (FOOOF) for each PSD.

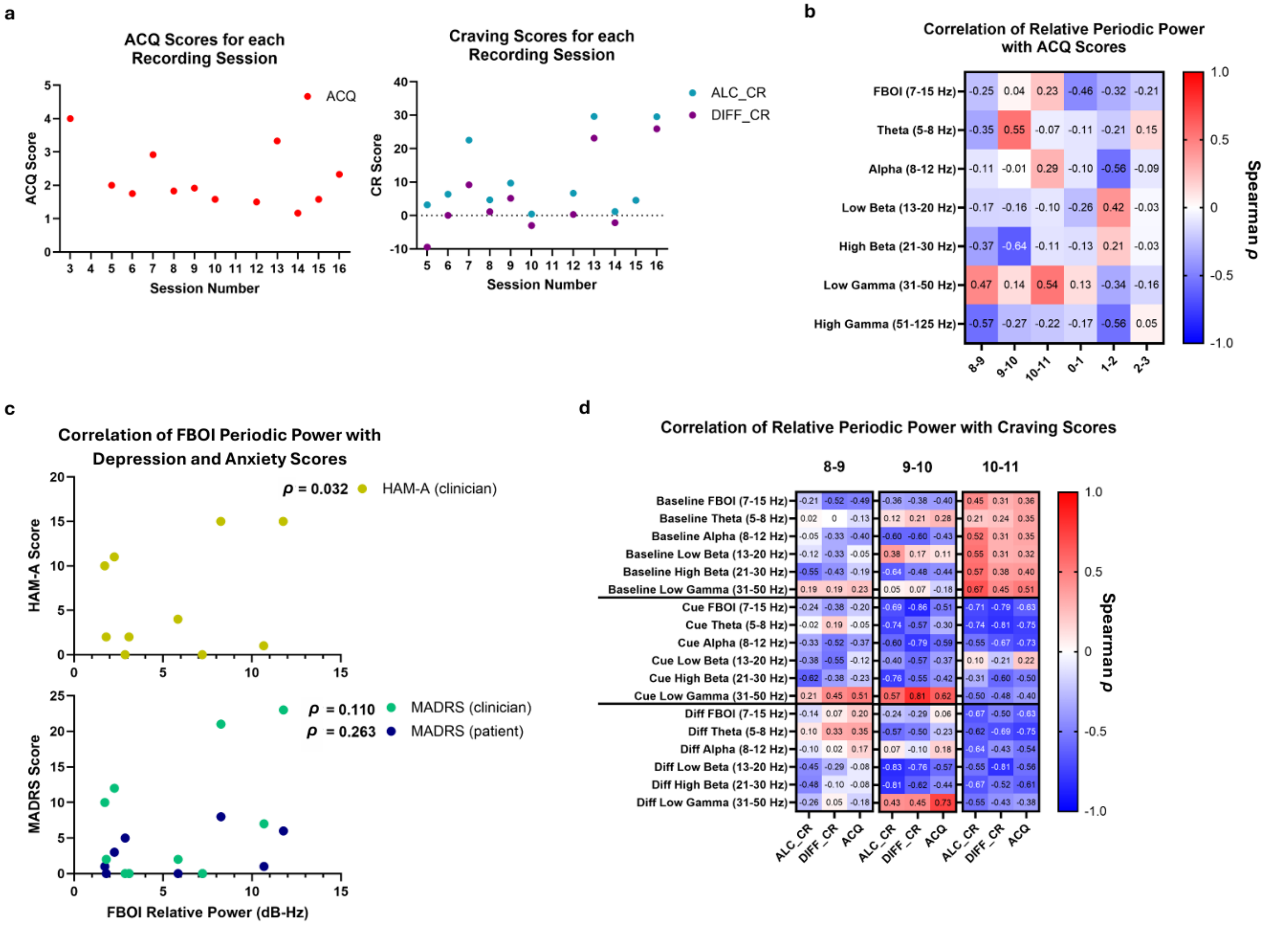


**Supplemental Figure 2. Craving scores did not show significant fluctuations or correlations with power of neural oscillations. (a)** Alcohol Craving Questionnaire (left, red), cue reactivity (CR) alcohol craving (right, blue), and CR differential craving (right, purple) scores displayed for each recording session. **(b)** Spearman's rank correlation coefficients for relative periodic powers (RPPs) from each sequential contact pair correlated to Alcohol Craving Questionnaire (ACQ) scores (*n* = 12, Šidák's multiple comparison test). **(c)** Scatterplots displaying the correlation between RPP from contact pair 8-9 across FBOI correlated to clinician HAM-A (top, yellow), clinician MADRS (bottom, light blue), and patient MADRS (bottom, dark blue) scores (*n* = 10, Spearman's rank correlation coefficient displayed). (**d**) Spearman's rank correlation coefficients for RPPs from contact pairs 8-9, 9-10, and 10-11 correlated to alcohol craving scores (*n* = 8, Šidák's multiple comparison test).

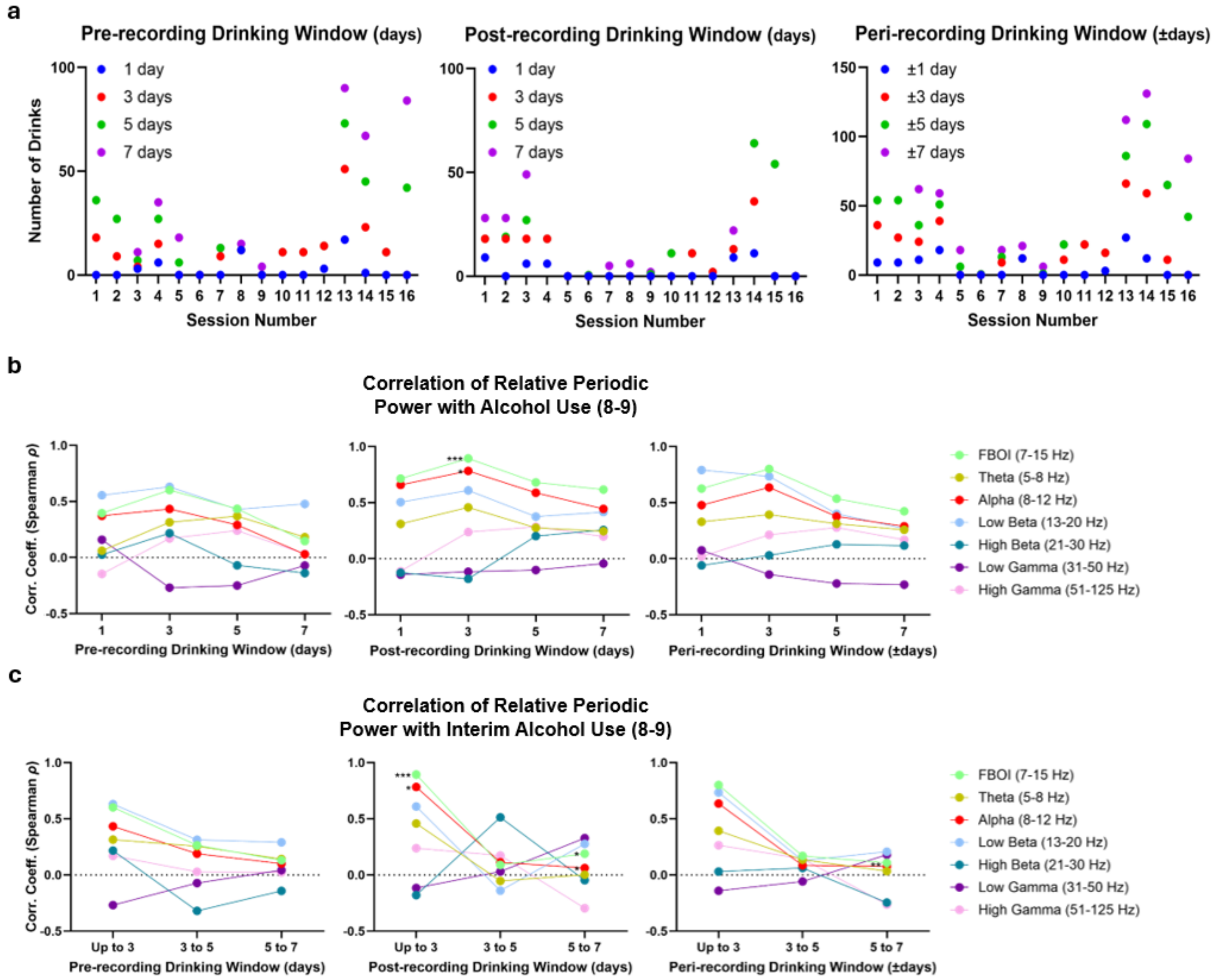


**Supplemental Figure 3. Interim alcohol use highlights higher correlations with post-recording session drinking windows. (a)** Total number of drinks post-, pre-, and peri-recording sessions displayed for each session. The drinking window refers to the number of days from the day of the recording that were used to determine the total number of drinks within that window. **(b)** Spearman's rank correlation coefficients for relative periodic powers (RPPs) from contact pair 8-9 correlated to total number of drinks post-recording session ($n = 16$). The drinking window refers to the number of days from the day of the recording that were used to determine the total number of drinks within that window (Šidák's multiple comparison test; ***$p < 0.001$, *$p < 0.05$). **(c)** Spearman's rank correlation coefficients for RPPs from contact pair 8-9 correlated to total number of drinks post-, pre-, and peri-recording sessions with interim alcohol use ($n = 16$). The drinking window refers to the number of days from the day of the recording that were used to determine the total number of drinks within that window (Šidák's multiple comparison test; ***$p < 0.001$, **$p < 0.01$, *$p < 0.05$).

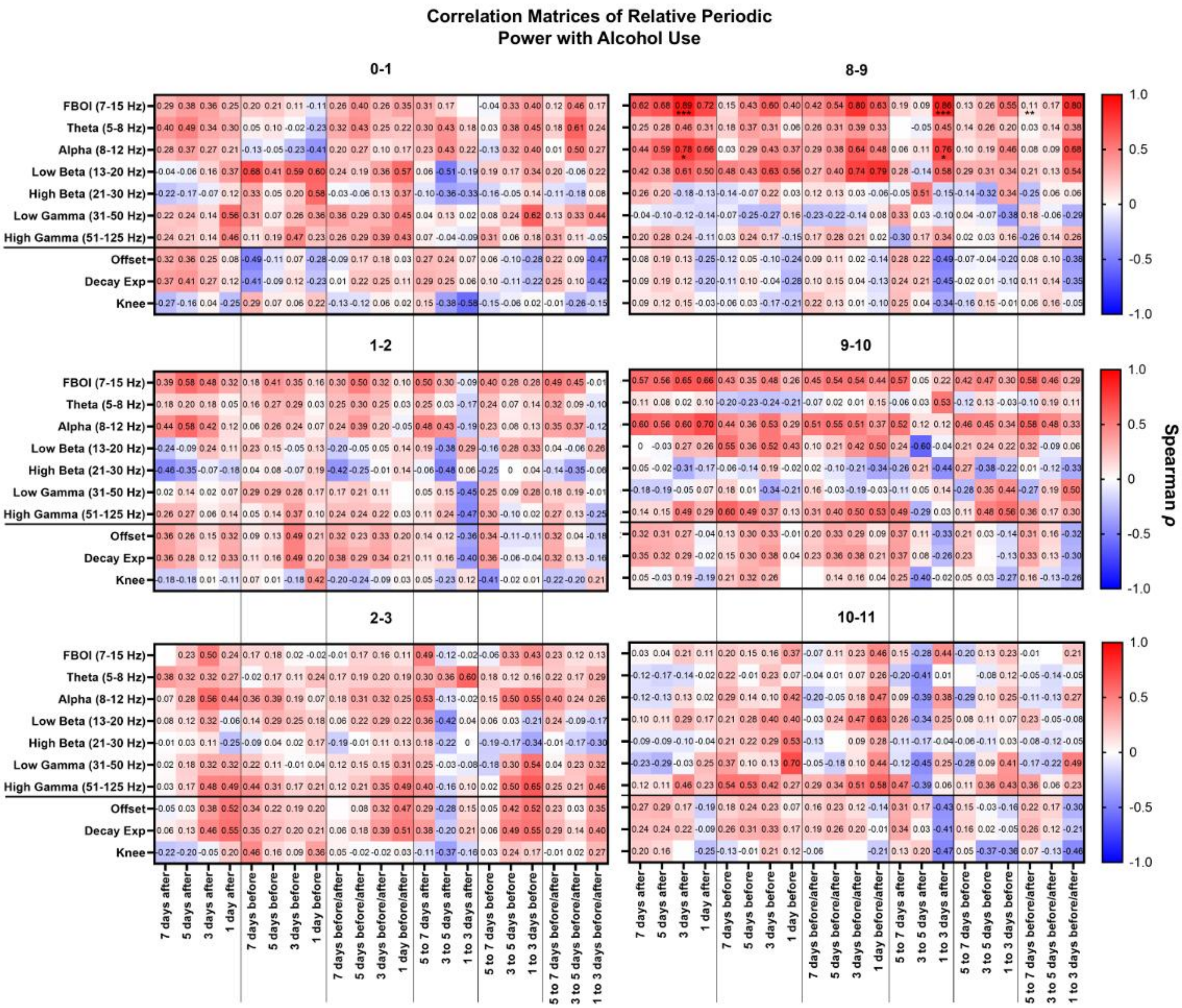


**Supplemental Figure 4. Correlation of RPP and aperiodic fit parameters with drinking behavior at the different DBS electrode contacts.** Spearman's rank correlation coefficients for relative periodic powers (RPPs) and aperiodic fit parameters (offset, decay exponent, and knee) from all sequential contact pairs correlated to total number of drinks post-, pre-, and peri-recording sessions as well as interim drinking windows. The drinking window refers to the number of days from the day of the recording that were used to determine the total number of drinks within that window (Šidák's multiple comparison test; ***$p < 0.001$, **$p < 0.01$, *$p < 0.05$).

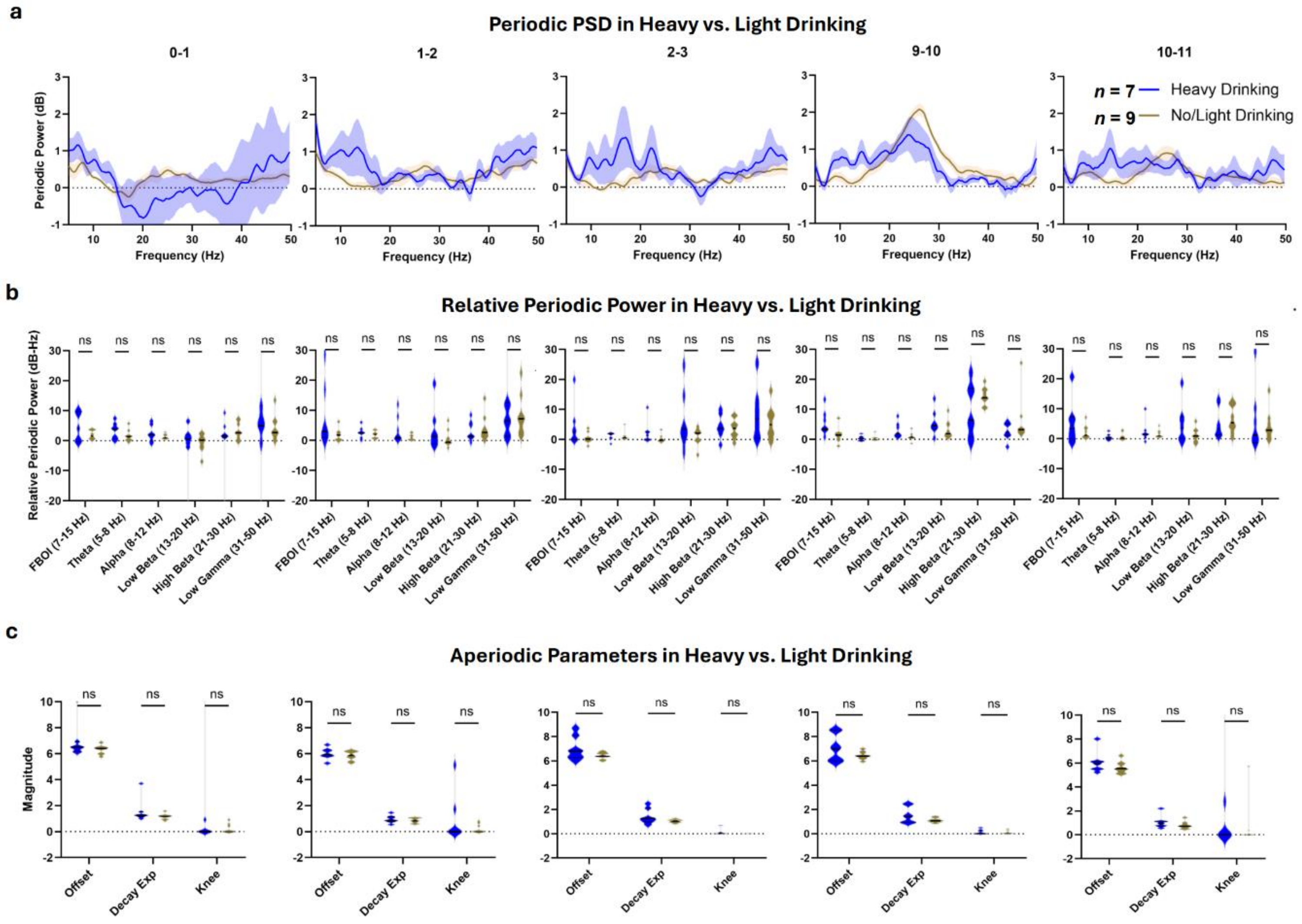


**Supplemental Figure 5. No significant relationship between heavy vs. light drinking and PSD power was observed other than at right most ventral electrode contacts (8-9).** (**a**) Mean periodic components of power spectral densities (PSDs) resulting from fitting of one over frequency (FOOOF) with standard error of the mean (translucent shading) grouped by heavy alcohol consumption following recording (blue, $n$ = 7) or no/light alcohol consumption following recording (brown, $n$ = 9). (**b**) Relative periodic power (shown with mean and standard error of the mean) in heavy drinking (blue) vs. no/light drinking (brown) (2-way repeated measures ANOVA, Šidák's multiple comparison test). (**c**) Aperiodic fit parameters compared in heavy drinking (blue) vs. no/light drinking (brown) with standard error of the mean shown (Mann-Whitney U test).
